# Spinal Movement Impairments in People with Acute Low Back Pain

**DOI:** 10.1101/2025.01.07.25320124

**Authors:** Kayla O. Krueger, Ryan P. Duncan, Linda R. van Dillen

## Abstract

**Objective:** Identify whether people with people with acute low back pain (LBP) display spinal movement impairments during clinical tests. Compare the prevalence of impairments in people with acute LBP to that of people with chronic LBP. Examine the effect on symptoms of systematically modifying impairments.

**Design:** Secondary analysis of data from a cross-sectional study on LBP.

**Methods:** 183 people with LBP were examined by a physical therapist using a standardized examination. Participants performed 9 primary tests using their preferred strategy. The clinician determined whether an impairment was present or absent. Participants reported the effect of the primary test on symptoms. If an impairment was present, it was modified during a secondary test. Participants reported the effect of the secondary test on symptoms relative to symptoms with the primary test. Chi-square tests of independence were used to test for differences in the proportion of impairments between people with acute LBP and people with chronic LBP. A McNemar-Bowker test was used to test whether there was a change in symptoms from the primary test to the secondary test.

**Results:** People with acute LBP displayed spinal movement impairments and the prevalence was similar to that of people with chronic LBP for 7 of the 9 primary tests. Most participants with symptomatic impairments reported their symptoms improved when the impairment was modified.

**Conclusion:** Spinal movement impairments are prevalent in people with acute LBP and may be modified during clinical tests to improve LBP symptoms.

## INTRODUCTION

Low back pain (LBP) is the leading cause of disability worldwide, impacting nearly 60-80% of adults in their lifetime.^4,6^ Non-specific LBP, defined as LBP not attributable to a known cause, represents the majority of all LBP cases.^1,10^ There is substantial variability in the clinical course of non-specific LBP.^9^ While some people recover quickly from an acute episode of LBP, up to 65% of people still report moderate pain and limitations in function at 12 months.^7^ Therefore, for many people, LBP becomes a long-term, function-limiting condition. Few factors consistently explain why some people recover quickly from an acute LBP episode and others transition to a recurrent or chronic course.^5^

How people with LBP move during functional activities may be one factor contributing to differences in the course of recovery from an acute LBP episode. Theoretical models such as the Kinesiopathologic Model provide a conceptual framework to understand the potential role of movement in the development and course of LBP.^17^ Based on the Model, LBP develops as a result of the repetitive use of the same patterns of movement during functional activities. The specific patterns of movement develop into spinal movement impairments (i.e., impairments of spinal movement control). Spinal movement impairments are characterized by the lumbar spine moving more readily into its available range of motion than other joints that can contribute to a movement goal.^14,18^ Over time, the repetitive use of the impairments during functional activities is proposed to contribute to altered loading of lumbar tissue, lumbar tissue stress, and eventually LBP and functional limitations.^2,15^

Spinal movement impairments have been found to be highly relevant to the clinical presentation and course of recovery in people with chronic LBP. Specifically, findings of prior work suggest the impairments are more prevalent in people with chronic LBP when compared to back-healthy people and are displayed across a series of clinical and functional tests.^11,12,14,18^ Moreover, the magnitude of the impairment has been associated with the intensity of a person’s symptoms and functional limitations.^13,14,28^ In many instances, treatment directed at modifying spinal movement impairments during clinical and functional tests has been shown to decrease symptoms and functional limitations in people with chronic LBP. ^14,19,26^

Given that spinal movement impairments are a potentially important, modifiable factor in chronic LBP, and an acute LBP episode precedes chronic LBP, examination of the role of these impairments in acute LBP is critical. Such an understanding could provide an important target for assessment and treatment from the outset of LBP. The primary purpose of the current study, therefore, was to conduct a secondary analysis of data collected during standardized clinical tests to identify whether people with acute LBP display spinal movement impairments. We hypothesized that people with acute LBP would display the impairments with the tests. We also compared the prevalence of spinal movement impairments in people with acute LBP to that of people with chronic LBP. We hypothesized that the prevalence of impairments with tests would be greater in people with chronic LBP compared to those with acute LBP. A secondary purpose was to examine the effect on symptoms of systematically modifying (i.e., improving) the impairments. We hypothesized that in the majority of people with acute LBP, modifying spinal movement impairments during symptomatic tests would result in an immediate improvement in symptoms.

## METHODS

### Participants

This is a secondary analysis of data collected from 183 people with non-specific LBP enrolled in a cross-sectional study examining the reliability and validity of a proposed movement -based classification system for LBP.^27,29^ Participants were recruited through advertisements in the St. Louis metropolitan area or from 1 of 6 outpatient physical therapy clinics where they were receiving treatment for their LBP. Inclusion and exclusion criteria for participants are provided in **TABLE 1**. All participants signed a written informed consent document approved by the Human Studies Committee of Washington University in St. Louis.

**TABLE 1.**
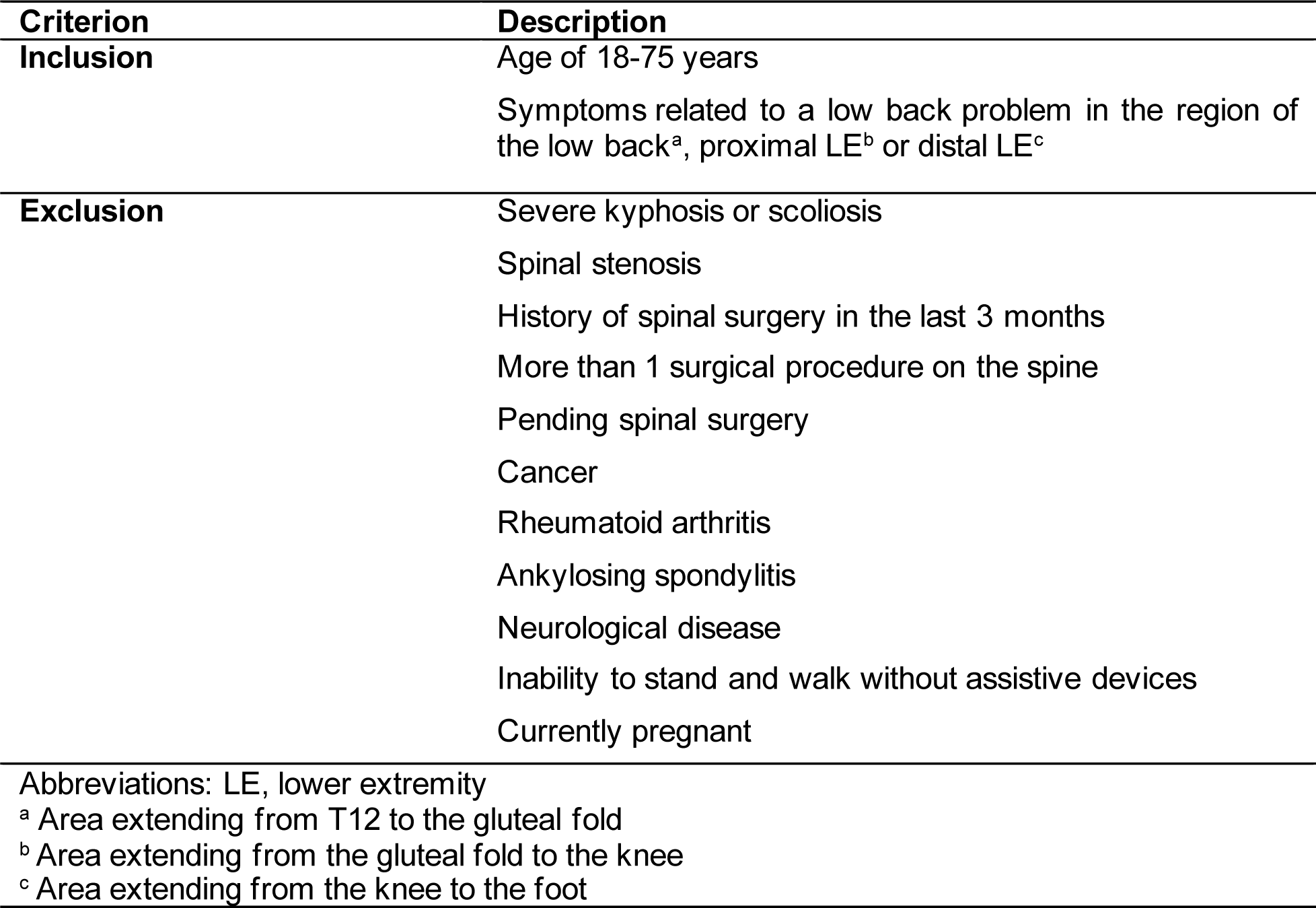
Inclusion and exclusion criteria for participants.

### Data Collection

Participants were examined by 1 of 5 reliably trained physical therapists using a standardized examination that included a history and physical examination.^27^ The history was obtained primarily though structured interview and consisted of questions related to demographics, medical and activity history, and LBP characteristics. Participants also completed a verbal descriptor scale that indexed the intensity of symptoms over the prior 7 days,^16^ and the Oswestry Low Back Pain Disability Questionnaire that indexed functional limitations.^3^

The physical examination included two types of movement tests. For the first type of test, referred to as the primary test, participants performed each of the following movements using their preferred strategy: (1) trunk forward bend, (2) trunk return from forward bend, (3) trunk lateral bend, (4) knee extension in sitting, (5) hip abduction and lateral rotation in supine, (6) knee flexion in prone, (7) hip rotation in prone, (8) rocking backward in quadruped, and (9) shoulder flexion in quadruped. Based on standardized criteria, a judgement was made by the therapist as to whether a spinal movement impairment was present or absent. Participants reported whether their symptoms increased, decreased, or remained the same during the primary test compared to a reference position. If an impairment was identified during a primary test, the test was immediately followed by a secondary test. During the secondary test the impairment was modified (i.e., improved), and the therapist assessed the effect of the modification on the participant’s symptoms. Secondary tests were performed for the following primary tests: (1) trunk forward bend, (2) trunk return from forward bend, (3) knee extension in sitting, (4) hip abduction and lateral rotation in supine, and (5) rocking backward in quadruped.^1^ The focus of the modifications was to alter the timing of movement of the lumbar spine relative to other joints, or the amount and type of lumbar spine and pelvic movement that occurred with each test. The examining therapist provided verbal instructions, demonstration, and physical assistance as needed to modify the impairment. Participants reported whether their symptomsincreased, decreased, or remained the same during the secondary test compared to the primary test. Definitions of symptom responses for primary tests and secondary tests are provided in **TABLE 2**. Descriptions of each primary and secondary test are provided in Van Dillen et al.^28^

**TABLE 2.** Definitions of symptom responses for primary tests and secondary tests.

| **Response** | **Definition** |
| --- | --- |
| **Increased** | The participant's symptoms were evoked, or increased in intensity, or were extended more distally |
| **Decreased** | The participant's symptoms were diminished or absent or were located more proximally relative to symptom responses with the reference test |
| **Remained the same** | The participant's symptoms were unchanged in intensity or location |

### Data Analysis

All analyses were performed using SPSS (IBM SPSS Statistics for Windows, Version 27.0 Armonk, NY, USA) with a two-tailed significance level set at p≤.05.

#### Participant characteristics

Participants were assigned to groups based on standardized definitions for acute LBP (i.e., symptom onset < 7 weeks) and chronic LBP. (i.e., symptom onset ≥ 7 weeks).^24^ Differences between participants with acute LBP and participants with chronic LBP in sex distribution, age, body mass index (BMI), number of LBP episodes in the prior 12 months, verbal descriptor scale values, and Oswestry Low Back Pain Disability Questionnaire scores were examined using a chi-square test of independence or an independent-groups t-test.

#### Prevalence of spinal movement impairments

For each of the 9 primary tests, the frequency of spinal movement impairments was calculated for people with acute LBP and people with chronic LBP. Chi-square tests of independence then were conducted for each primary test to test for differences in the proportion of impairments between the two groups. For primary tests that were performed with each limb (i.e., knee extension in sitting, hip abduction and lateral rotation in supine, hip rotation in prone, shoulder flexion in quadruped), an impairment was recorded as present if it was observed with 1 or both limbs. Fisher’s Exact test was used when the expected values assumption was not met. *Symptoms with movement*

The analysis included data from people with acute LBP that demonstrated an impairment during a primary test. Only primary tests that were followed by a secondary test were analyzed (n=5). The frequency of symptom responses (i.e., increased, decreased, remained the same) was calculated for both the primary and secondary tests. McNemar-Bowker tests then were performed to test whether there was a change in symptoms from the primary test to the secondary test. *A priori* pairwise comparisons were examined using a McNemar test when the test statistic was significant. A binomial Exact test was used when the expected values assumption was not met.

## RESULTS

### Participant characteristics

The sample included 183 participants. Fifty-one participants had acute LBP and 132 had chronic LBP. There were no differences between the groups other than the chronic LBP group was older (p=.04) and heavier (p<.01) than the acute LBP group (**TABLE 3**).

**TABLE 3.**
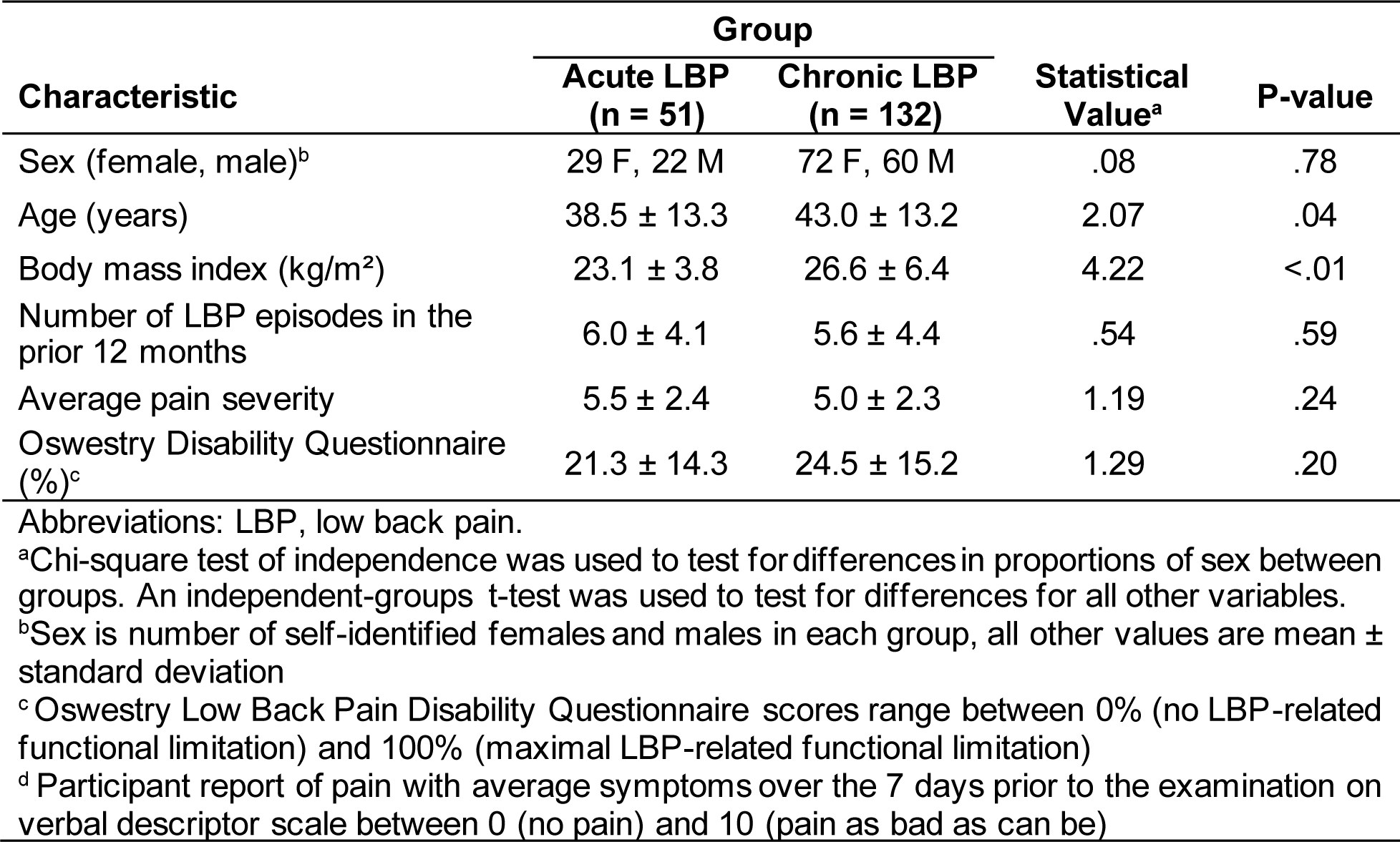
Participant characteristics.

| **Characteristic** | **Group** |  | **Statistical Value<sup>a</sup>** | **P-value** |
| --- | --- | --- | --- | --- |
|  | **Acute LBP**<br>**(n = 51)** | **Chronic LBP**<br>**(n = 132)** |  |  |
| Sex (female, male)<sup>b</sup> | 29 F, 22 M | 72 F, 60 M | .08 | .78 |
| Age (years) | 38.5 ± 13.3 | 43.0 ± 13.2 | 2.07 | .04 |
| Body mass index (kg/m<sup>2</sup>) | 23.1 ± 3.8 | 26.6 ± 6.4 | 4.22 | <.01 |
| Number of LBP episodes in the prior 12 months | 6.0 ± 4.1 | 5.6 ± 4.4 | .54 | .59 |
| Average pain severity | 5.5 ± 2.4 | 5.0 ± 2.3 | 1.19 | .24 |
| Oswestry Disability Questionnaire (%)<sup>c</sup> | 21.3 ± 14.3 | 24.5 ± 15.2 | 1.29 | .20 |

Abbreviations: LBP, low back pain.
<sup>a</sup>Chi-square test of independence was used to test for differences in proportions of sex between groups. An independent-groups t-test was used to test for differences for all other variables.
<sup>b</sup>Sex is number of self-identified females and males in each group, all other values are mean ± standard deviation
<sup>c</sup>Oswestry Low Back Pain Disability Questionnaire scores range between 0% (no LBP-related functional limitation) and 100% (maximal LBP-related functional limitation)
<sup>d</sup>Participant report of pain with average symptoms over the 7 days prior to the examination on verbal descriptor scale between 0 (no pain) and 10 (pain as bad as can be)

### Prevalence of spinal movement impairments

There were no differences in the proportions of impairments between people with acute LBP and people with chronic LBP for 7 of the 9 primary tests. Compared to people with acute LBP, a greater proportion of people with chronic LBP displayed an impairment during knee extension in sitting (X^2^=3.70, p=.05) and knee flexion in prone (X^2^=7.41, p<.01; **TABLE 4**).

**TABLE 4.**
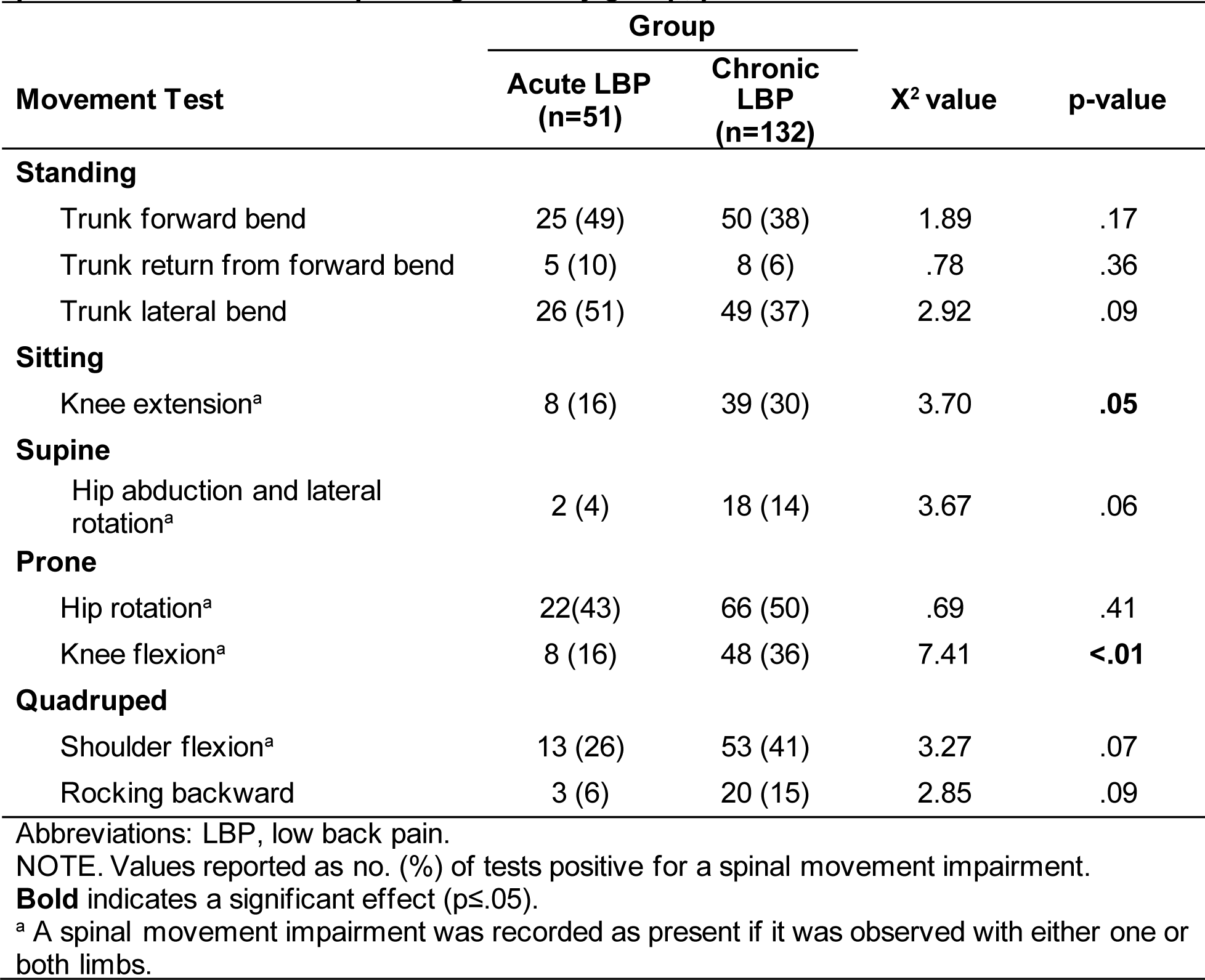
Prevalence of spinal movement impairments in people with acute low back pain or chronic low back pain organized by group, position and test.

### Symptoms with movement

A McNemar-Bowker test was performed for 1 of the 5 movement tests, i.e., trunk forward bend. There was a significant change in symptoms from the primary test to the secondary test (X^2^=14, p=.003; **TABLE 5**). Post-hoc analyses revealed that a significant number of people who reported increased symptoms during the primary test reported decreased symptoms during the secondary test. All other post-hoc comparisons were not significant. Results are reported qualitatively for the other 4 tests because the assumptions of the McNemar-Bowker test were not met. For each of the 4 tests, the majority of people who reported increased symptoms during the primary test reported decreased symptoms during the secondary test (**TABLE 5**).

**TABLE 5.** Symptom responses reported for each primary test and secondary test for people with acute LBP that demonstrated a spinal movement impairment.

| Movement Test |  |  | Secondary Test |  |  |  |
| --- | --- | --- | --- | --- | --- | --- |
|  |  |  | Decreased | Same | Increased | Total |
| Trunk forward bend<sup>a</sup> | Primary Test | Decreased | 1 | 2 | 0 | 3 |
|  |  | Same | 2 | 5 | 0 | 7 |
|  |  | Increased | 13 | 1 | 1 | 15 |
|  |  | Total | 16 | 8 | 1 | 25 |
| Trunk return from forward bend<sup>b</sup> | Primary Test | Same | 0 | 1 | 0 | 1 |
|  |  | Increased | 3 | 0 | 1 | 4 |
|  |  | Total | 3 | 1 | 1 | 5 |
| Knee extension<sup>b</sup> | Primary Test | Same | 0 | 4 | 0 | 4 |
|  |  | Increased | 4 | 0 | 0 | 4 |
|  |  | Total | 4 | 4 | 0 | 8 |
| Hip abduction and lateral rotation<sup>b</sup> | Primary Test | Same | 0 | 1 | 0 | 1 |
|  |  | Increased | 1 | 0 | 0 | 1 |
|  |  | Total | 1 | 1 | 0 | 2 |
| Rocking backward<sup>b</sup> | Primary Test | Same | 0 | 1 | 0 | 1 |
|  |  | Increased | 2 | 0 | 0 | 2 |
|  |  | Total | 2 | 1 | 0 | 3 |

Abbreviations: LBP, low back pain.
<sup>a</sup> Movement test for which McNemar-Bowker chi-square test was calculated ( $X^2 = 14$ , $p = .003$ )
<sup>b</sup> Tests in which no person reported a decrease in symptoms with the primary test

## DISCUSSION

Our purpose was to identify whether people with acute LBP displayed spinal movement impairments reported to be important in people with chronic LBP. People with acute LBP displayed the impairments across 9 standardized clinical movement tests. We also compared the prevalence of spinal movement impairments in people with acute LBP to that of people with chronic LBP for each of the 9 tests. The prevalence between groups was similar for the majority (7/9; 78%) of the tests examined. Finally, in people with acute LBP who displayed an impairment during a primary test we examined the effect on the person’s symptoms of a secondary test to modify the impairment. In the majority of people, modifying impairments that were symptomatic resulted in an immediate improvement in symptoms.

A recent clinical commentary suggests there are differences in spinal movement control between people with LBP and back-healthy people. It is proposed that the differences can be characterized along a spectrum defined by “tight” control and “loose” control strategies. “Tight” control is characterized by restricted movement of the spine and is associated with increased trunk muscle activation and stiffness. Alternatively, “loose” control is characterized by faster and larger amplitude movements of the spine, and is associated with reduced trunk muscle activation and stiffness.^25^ Both ends of the spectrum may have detrimental effects on spine health. “Tight” control may lead to reduced movement variability and increased compressive loading of spinal tissue whereas “loose” control may lead to excessive shear forces and tissue strains.

Findings from our prior work suggest many people with chronic LBP adopt a “loose” control strategy. These findings are evidenced in studies comparing early-phase movement of the lumbar spine between people with chronic LBP and back-healthy people. In one study, people with chronic LBP demonstrated earlier lumbopelvic motion than back-healthy people during 2 different clinical tests, i.e., loose control.^18^ Another study compared movement between people with chronic LBP and back-healthy people during a functional activity test of picking up an object. People with chronic LBP demonstrated a greater magnitude of lumbar flexion in the first half of the descent phase compared to back-healthy people, i.e., loose control.^14^

The few studies examining spinal movement control in people with acute LBP suggest that people with acute LBP tend to adopt a “tight” control strategy. In one study, Shojaei et al^21^ reported that the relative contribution of lumbar flexion to total movement during forward bend was smaller in women with acute LBP (LBP ≤ 3 months) compared to back-healthy women, i.e., tight control. Consistent with these findings, Shum et al^22,23^ reported that the relative contribution of lumbar flexion to total movement during a series of functional activity tests was smaller in men with acute LBP compared to back-healthy men, i.e., tight control.

In the current study, many people with acute LBP display spinal movement impairments similar to the impairments we have identified in people with chronic LBP, i.e., a “loose” control strategy. Thus, our findings are inconsistent with what has been reported in some previous studies. One potential reason for the discrepancy is that the type of variable used to quantify spinal movement control differs between the current study and prior work. In prior work, spinal movement control is most often indexed as the relative contribution of the lumbar spine to total movement at end of the range of the test motion. Whereas the spinal movement impairments of interest in the current study examine excursion of the lumbar spine in the first half of the test motion or the timing of movement of the lumbar spine relative to other joints during the test motion. We focus on this index of spinal movement control in people with acute LBP because the impairments have been found to be a potentially important, modifiable factor in people with chronic LBP.^14,19,26^

A more recent study compared movement control of the trunk and pelvis in the frontal plane in people with recurrent LBP and back-healthy people during gait. To examine the effect of acute pain on spinal movement control, people with recurrent LBP were tested twice, first, when they were in an acute episode of LBP, and then when their pain was in remission. This within-person repeated measures design offers an advantage over previous cross-sectional studies. Instead of comparing people with LBP to back-healthy people, this design focuses on changes within the same person. This approach reduces the amount of between-subject variability, thereby providing a better estimate of the effect of acute pain on movement. The researchers found that people with recurrent LBP, regardless of pain status, demonstrated a “loose” control strategy compared to back-healthy people. Interestingly, comparisons within the recurrent LBP group revealed that during the remission condition, people were even more “loose” than in the acute pain condition.^20^ The findings from this study offer partial support for our findings of the use of a “loose” control strategy in some people with acute LBP.

We demonstrated that systematically modifying the impairment with a secondary test resulted in an immediate improvement in LBP in the majority of people with acute LBP. In a few cases people reported that the modifications with the secondary test did not change (n=15 occurrences) or increased symptoms (n=2 occurrences) they experienced during the primary test. There are potential reasons for these responses with a secondary test for some people. First, the presence of increased muscle activation and trunk stiffness may have been present during the secondary test making it difficult to adequately modify the impairment. Second, pre-existing postural conditions may have made it difficult to modify the impairment. For example, some participants presented with conditions such as kyphosis (n=3), swayback (n=1), and increased flexion of the lumbar curve (n=12) which may have made the impairment non-modifiable or non-modifiable within one session. Third, there may be other non-movement related factors, such as central sensitization, contributing to a person’s perception of LBP.^8^ Because examiners were not required to assess other potential contributors to LBP, the extent to which these factors were present is unknown.

Our findings are important because we have shown that spinal movement impairments found to be associated with symptoms and functional limitations in people with chronic LBP are present in some people with acute LBP. Moreover, for the majority of people with acute LBP, the impairments can be modified to improve symptoms. These results suggest that the spinal movement impairments described may be an important factor to examine in more depth in people with acute LBP. An important next step would be to measure the impairments more precisely using robust methodology (e.g., motion-capture, movement sensors) and to determine whether the impairments are associated with symptom intensity and functional limitations.

Our study has limitations. First, compared to the acute LBP group, the chronic LBP group was significantly older (difference = 4.5 years; p=0.04) and heavier (BMI difference = 3.5 kg/m^2^; p<.01). Although statistically significant, the magnitude of the differences is small and thus unlikely to have a large impact on findings. Second, for movement tests performed on both limbs (e.g., knee flexion performed in prone), an impairment was recorded as present if observed with either one or both limbs. Therefore, the number of impairments during tests of limb movements in each group may be underestimated. Finally, the presence of an impairment was based on clinician judgement. Clinicians, however, used standardized criteria and reliability of judgments during these clinical tests has been found to be acceptable.^27^ In addition, in any instance in which a clinician was uncertain if an impairment was present, the default was to record that there was no impairment present. Therefore, the decision rule potentially would contribute to an underestimate of prevalence.

## CONCLUSION

Our findings suggest that some people with acute LBP display spinal movement impairments similar to people with chronic LBP, and that the impairments appear to be relevant to a person’s LBP symptoms. Given the importance of spinal movement impairments in people with chronic LBP, further examination of the role of the impairments in people with acute LBP is warranted.

## KEY POINTS

- **Findings:** People with acute LBP displayed spinal movement impairments during clinical tests and the prevalence was similar to that of people with chronic LBP for most tests. Modifying symptomatic impairments led to an improvement in symptoms in the majority of people with acute LBP.
- **Implications:** Spinal movement impairments could be an important target for assessment and treatment in people with acute LBP.
- **Caution:** The presence of spinal movement impairments was based on clinician judgement.

## Study Details

### Ethical Approval

The study was approved for secondary analysis by the Washington University School of Medicine Institutional Review Board (IRB#: 202102112).

### Authors’ Contributions Statement

Concept/idea/research design: KK, LVD, RD. Investigation: KK, LVD, RD. Analysis and interpretation of data: KK, LVD, RD. Writing/review/editing of manuscript: KK, LVD, RD. Final approval of the manuscript: KK, LVD, RD

### Data Sharing Statement

All data relevant to the study are included in the manuscript or are available as supplementary files.

### Patient and Public Involvement

Study participants and/or the public were not involved in the study design, data collection, data analysis, data interpretation, or writing of the current research.

## Footnotes

1 At the time of the original study secondary tests were developed and standardized for 5/9 primary tests.

## Notes

### Competing Interest Statement

The authors have declared no competing interest.

### Funding Statement

This research received funding from the Foundation for Physical Therapy (Grant No. 94R-03-N0R-02), KK is supported by a training grant from National Institutes of Health (Grant No. T32DH007434). The funders of the study had no other role in the study design, data collection, data analysis, data interpretation, or writing of the current research. KK reports grant funding from the NIH during conduct of the study. RD reports grant funding from the NIH during conduct of the study. LVD reports grant funding from the NIH and from the Academy of Orthopaedic Physical Therapy during conduct of the study. No other disclosures were reported.

### Author Declarations

The Washington University School of Medicine Institutional Review Board (IRB#: 202102112) gave ethical approval for secondary analysis of the study.

### Summary of Updates

Table 3 statistical value/p-value is revised.

